# Developing a User-Friendly Oropharyngeal HPV-16 Point-of-Care Diagnostic: HPV-16 Virus Detection in Oral Rinse Sample Matrix With Colorimetric Loop-Mediated Isothermal Amplification

**DOI:** 10.1101/2024.05.15.24307441

**Authors:** John Halphen, Nicholas A. Wright, Samantha Fisher, San Juana I. Juarez, Lisa Sanchez, Timothy E. Riedel

## Abstract

Human papillomavirus type 16 (HPV-16) is a highly carcinogenic variant that is responsible for the majority of human oropharyngeal cancers. HPV-16 is critically under-tested, and there are currently no commercially available HPV-16 diagnostics for oral infection sites. We developed an oral HPV-16 diagnostic system with the goal of being as user-friendly and non-invasive as possible. The system tests an oral rinse sample from the patient, is interpreted via a color change from pink to yellow, and can be incubated in a battery-powered coffee mug. The colorimetric loop-mediated isothermal amplification-based diagnostic is sensitive to synthetic HPV-16 E7/E1 DNA at levels as low as 10^3^ copies after 20 minutes of incubation. This study supports the potential to develop an easy-to-assemble and easy-to-use diagnostic for oropharyngeal HPV-16 infections.

## 1. Introduction

Human papillomavirus (HPV) is a sexually transmitted disease classified as a carcinogen, meaning that infection correlates with the prevalence of certain types of cancer (Li et al., 2017). Of the fourteen high-risk variants, HPV-16 and 18 have been identified as the strains responsible for the highest prevalence of several types of cancer (Siegler et al., 2019). HPV is associated with 63% of oropharyngeal cancers, and of HPV-related oropharyngeal cancers, 95% of those cases coincide with HPV-16 infection (Li et al., 2017). Currently, there are no diagnostics approved by the FDA for the detection of HPV in sites of infection other than the cervix (Vives et al., 2020). This is particularly problematic due to the uniquely localized nature of HPV infection and subsequent carcinogenic activity, primarily acting at the site of initial infection (Lyford-Pike et al., 2013). Therefore, testing the genitals is ineffectual in diagnosing HPV if the site of the initial infection is the oral cavity.

Loop-mediated isothermal amplification (LAMP) is a nucleic acid amplification system (Notomi et al., 2000) that has the potential to be more user-friendly than PCR-type reactions (Li et al., 2022). LAMP-based tests can be completed outside of a laboratory since no thermocycler is required, and they can be set up to have a simple readout (Notomi et al., 2015). In this study, we explore the feasibility of a simple-to-use diagnostic for HPV-16 infection of the oropharyngeal site by characterizing the sensitivity of LAMP combined with a pH indicator when inoculated with an oral rinse sample and incubated in a battery-powered coffee mug.

## 2. Materials and Methods

### 2.1. Oral rinse and saliva preparation

Oral rinse experiments were prepared using commercially available bottled water (Aquafina, 16.9 oz). Oral rinses were collected and pooled from four participants who abstained from eating food for at least 30 minutes and drinking water for at least 5 minutes before collection. Each participant rinsed and gargled with 15 mL of bottled water (pre-poured into a disposable plastic cup) for 30 seconds before spitting the contents of their mouth back into the cup (D’Souza et al., 2005). Equal proportions of participant rinses were mixed to yield a de-identified oral rinse pool. This oral rinse pool was used in all reported LAMP reactions that utilized an oral rinse sample matrix. The saliva collection protocol described was reviewed and approved as ethical (IRB Protocol 2013-12-007).

The saliva sample matrix was prepared by having four participants fill a centrifuge tube with 2.5 mL of saliva following the same pre-collection protocol described above. The saliva was then sampled from and mixed in equal proportions in another centrifuge tube to yield a de-identified saliva pool. This pooled saliva was added to the LAMP reactions that used saliva as the sample matrix.

### 2.2. Colorimetric LAMP assay construction

We utilized a colorimetric LAMP system (New England BioLabs, Ipswich MA, USA, cat. #M1800) as a qualitative measure for the amplification of the target DNA sequence. These reactions change color from pink to yellow due to pH change resulting from DNA amplification. Previously published LAMP primers targeting the HPV-16 E7/E1 genes (Saetiew et al., 2011, Table 1) were used in all reactions.

**Table 1.**
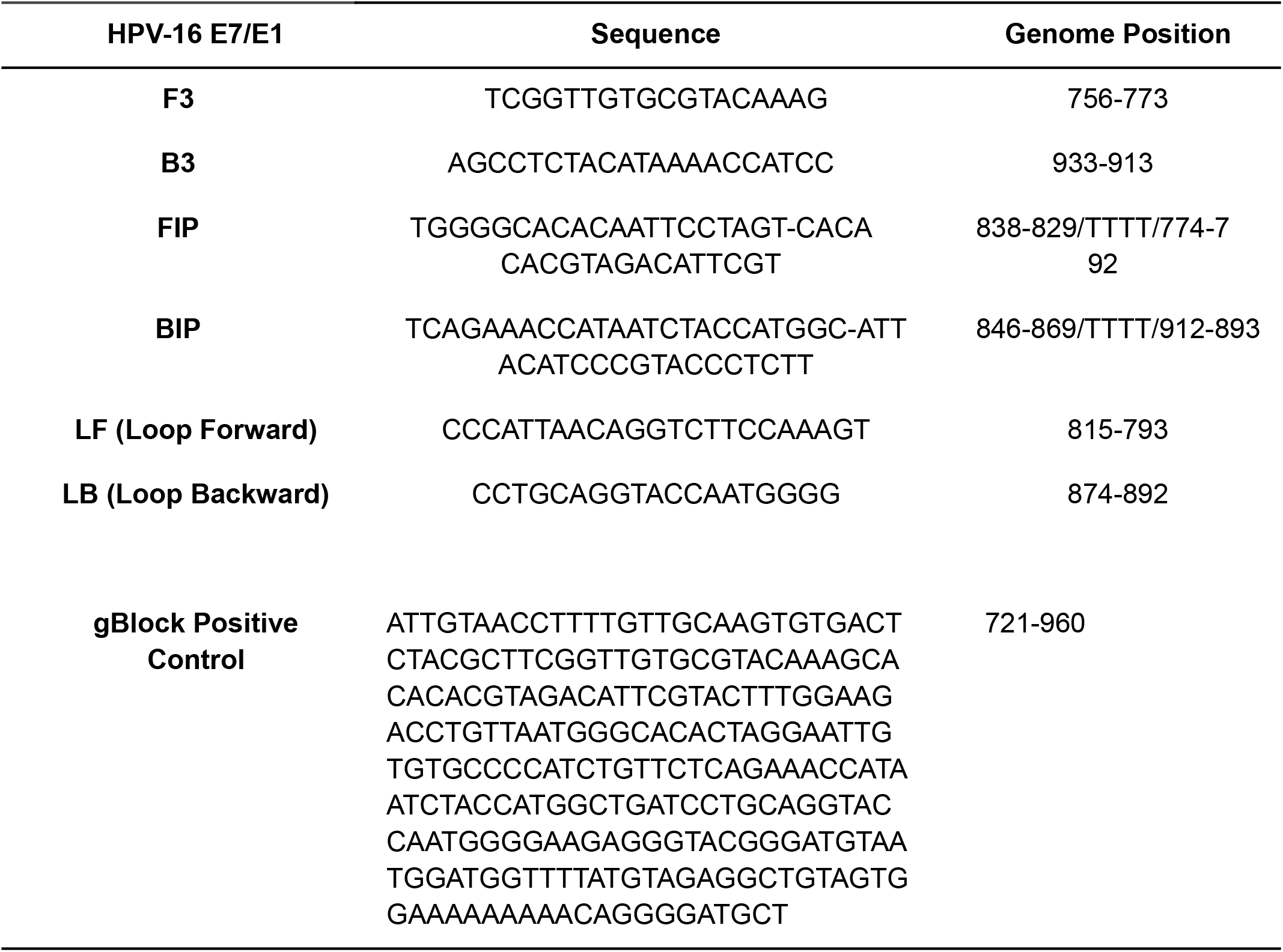
LAMP primers and positive control HPV-16 E7/E1 sequence (Saetiew et al., 2011) used in this study are reported in the 5’ to 3’ direction.

In 0.2 mL tubes (VWR PCR Tubes #53509-304) 25 μL reactions were constructed with the following: 12 ul of NEB 2X Colorimetric Master Mix (contains all necessary ingredients for colorimetric LAMP except primers and template), 2.5 μl of a 10X mix of all primers for a final concentration of 0.2 μM F3 and B3, 1.6 μM FIP and BIP, and 0.4 μM Loop Forward and Loop Backward (Table 2).

**Table 2.** Storage and reaction LAMP primer concentrations used for this study.

| **Primer** | **10X Stock ($\mu\text{M}$)** | **1X Reaction Concentration ($\mu\text{M}$)** |
| --- | --- | --- |
| **F3** | 2 | 0.2 |
| **B3** | 2 | 0.2 |
| **FIP** | 16 | 1.6 |
| **BIP** | 16 | 1.6 |
| **LF (Loop Forward)** | 4 | 0.4 |
| **LB (Loop Backward)** | 4 | 0.4 |

These components were combined on the bench at room temperature with an additional 10.5 μl of sample matrix (molecular grade water, oral rise, or saliva) added to bring the volume up to 24 μl. The final microliter was used to inoculate reactions with either molecular biology grade water (Corning, cat. # 46-000-CM) for non-template negative controls (NTC) or with 10^3^ copies of the HPV-16 E7/E1 DNA for positive template controls (PTC).

PTC synthetic dsDNA was purchased (gBlock, Integrated DNA Technologies, Inc.), spanning part of the HPV-16 E7/E1 genes (Saetiew et al., 2011). The dsDNA was resuspended with TE buffer to a concentration of 10^10^ copies per μL. and stored frozen at -20 °C. On the same day as the experimental runs, a serial dilution was performed on the thawed 10^10^ copies per μL stock with molecular water to a final 10^3^ copies per μL concentration.

### 2.4. Incubating the LAMP reactions and scoring results

An 8-strip of PCR tubes containing the reaction mixture was incubated in either a laboratory water bath set to 65 °C or an Ember Temperature Control Smart Mug 2 (EM2), a battery-powered temperature-controlling coffee mug (Ember Technologies, Inc., 14oz #CM19P), set to its highest setpoint of 62.5 °C with 150 mL of water preheated using the laboratory water bath. All EM2 incubations were completed with the EM2 sitting on the saucer charger system.

To promote even heating of the reactions and extend battery life, the EM2 was operated with a styrofoam cap over the top opening. Reactions were scored every 10 minutes during incubation by removing the tubes from the incubator before photographing and returning them to the incubator, taking approximately 15 seconds. Reaction colors were scored with the unaided eye as either pink (P), orange (O), yellow (Y), or with color combinations to represent intermediate levels when needed (e.g., O/Y for an orange-yellow mixture).

## 3. Results

### 3.1. Validation of the Saetiew et al. 2011 LAMP primers

We tested the described HPV16 E7/E1 LAMP reaction with molecular water (molH_2_O) as the sample matrix to validate the Saetiew et al. 2011 primers (Table 1) in combination with the colorimetric LAMP reaction mix (Section 2.2). PTCs were inoculated with 10^4^ copies of HPV16 E7/E1 DNA and NTCs were inoculated with molH_2_O with no added DNA. These LAMP reactions were scored for amplification (Section 2.4) every 10 minutes over 70 minutes of incubation at 65 °C (Table 3, Figure 1).

**Table 3:** Validation of Saetiew et al. 2011 primers with the NEB colorimetric LAMP system. PTC molH_2_O reactions spiked with 10^4^ copies of HPV16 E7/E1 DNA and NTC reactions with no added DNA were scored (P represents no DNA amplification, O represents some DNA amplification, Y represents full DNA amplification, and color combinations represent intermediate levels) every 10 minutes of incubation in a water bath at 65 °C.

|  | 10 min | 20 min | 30 min | 40 min | 50 min | 60 min | 70 min |
| --- | --- | --- | --- | --- | --- | --- | --- |
| NTC-1 | P | P | P | P | P | P | P |
| NTC-2 | P | P | P | P | P | P | O |
| NTC-3 | P | P | P | P | P | P | P |
| PTC-1 | P | P | Y | Y | Y | Y | Y |
| PTC-2 | P | P | Y | Y | Y | Y | Y |
| PTC-3 | P | P | Y | Y | Y | Y | Y |

**Figure 1:**
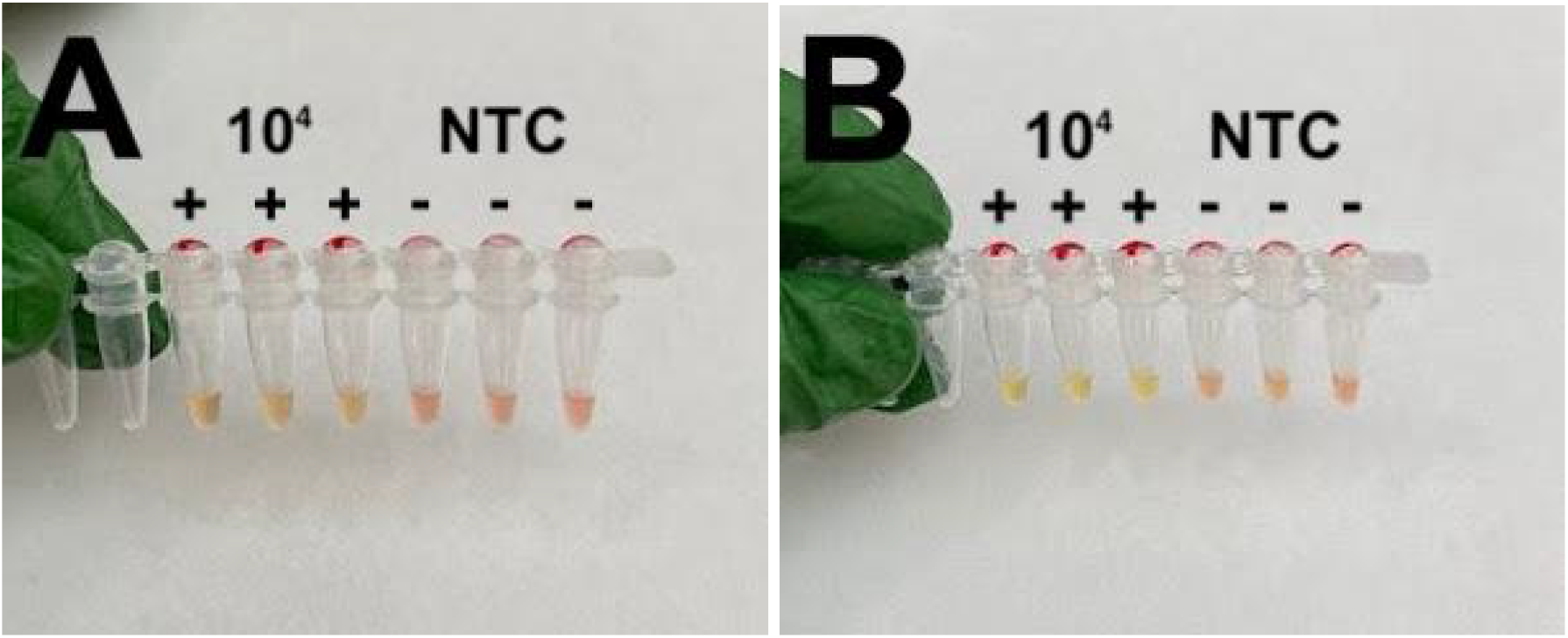
Validation of Saetiew et al. 2011 primers with the NEB colorimetric LAMP system. PTC molH_2_O reactions spiked with 10^4^ copies of HPV16 E7/E1 DNA (indicated by + symbols) and NTC reactions with no added DNA (indicated by - symbols) after 30 minutes (A) and 70 minutes (B) of incubation in a water bath at 65 °C.

The assays were found to be sensitive to 10^4^ copies of HPV16 E7/E1 DNA with full amplification of the PTCs by 30 minutes and no indication of NTC amplification for at least 60 minutes.

### 3.2. Impact of saliva or oral rinse on LAMP performance

After testing the Saetiew et al. 2011 LAMP primers with the NEB 2X Colorimetric Master Mix (Section 3.2), we tested de-identified pooled saliva and oral rinse as the sample matrix of the assay. These reactions were assembled in the same fashion as before but with molH2O substituted with either saliva or oral rinse (Section 2.1). Reactions were incubated in a water bath for 70 minutes and scored for color change every 10 minutes (Tables 4 and 5, Figure 2).

**Table 4:** Testing saliva as the sample matrix. PTC saliva reactions spiked with 10^4^ copies of HPV16 E7/E1 DNA and NTC saliva reactions with no added DNA were scored (P represents no DNA amplification, O represents some DNA amplification, Y represents full DNA amplification, and color combinations represent intermediate levels) every 10 minutes of incubation in a water bath at 65 °C.

|  | 10 min | 20 min | 30 min | 40 min | 50 min | 60 min | 70 min |
| --- | --- | --- | --- | --- | --- | --- | --- |
| NTC-1 | P | P | P | P | P | P | P/O |
| NTC-2 | P/O | O | O | O | O | O | O/Y |
| PTC-1 | P | P/O | O | O | O/Y | O/Y | O/Y |
| PTC-2 | P | P/O | O | O | O/Y | O/Y | O/Y |

**Table 5:** Testing oral rinse as the sample matrix. PTC oral rinse reactions spiked with 10^4^ copies of HPV16 E7/E1 DNA and NTC oral rinse reactions with no added DNA were scored (P represents no DNA amplification, O represents some DNA amplification, Y represents full DNA amplification, and color combinations represent intermediate levels) every 10 minutes of incubation in a water bath at 65 °C.

|  | 10 min | 20 min | 30 min | 40 min | 50 min | 60 min | 70 min |
| --- | --- | --- | --- | --- | --- | --- | --- |
| NTC-1 | P | P | P | P | P | P | P/O |
| NTC-2 | P | P | P | P | P | P | P/O |
| PTC-1 | P/O | P/O | O | Y | Y | Y | Y |
| PTC-2 | P/O | P/O | O | Y | Y | Y | Y |

**Figure 2:**
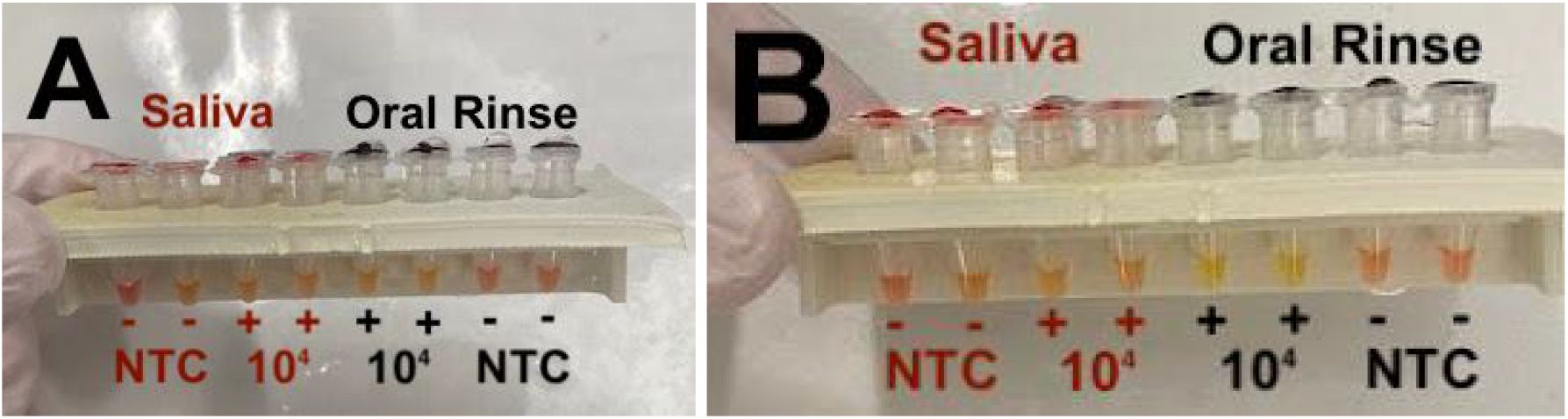
Testing saliva and oral rinse as the sample matrix. PTC saliva (red labels) or oral rinse (black labels) reactions spiked with 10^4^ copies of HPV16 E7/E1 DNA (indicated by + symbols) and NTC reactions with no added DNA (indicated by - symbols) after 40 minutes (A) and 70 minutes (B) of incubation in a water bath at 65 °C.

The LAMP assay using oral rinse as the sample matrix performed similarly to the assay using molH2O, both having their PTC reactions undergo amplification and color change after 30 minutes, while their NTC reactions changed color only after 70 minutes of incubation at 65 °C in the water bath. The assay using pooled saliva as the sample matrix had PTC reactions that were not distinguishable from NTC reactions regarding color change timing. We ran several more tests of 1/2 and 1/4 dilutions of the pooled saliva by molH2O. We found that both PTCs and NTCs changed colors from pink to yellow after 30 minutes (data not shown).

### 3.3. Impact of EM2 Mug incubation on LAMP performance

With the saliva not working well, we did not carry out further saliva-based tests. We further characterized the oral rinse colorimetric LAMP system by testing its performance with more dilute inocula and incubation in the EM2 mug (Figure 3).

**Figure 3:**
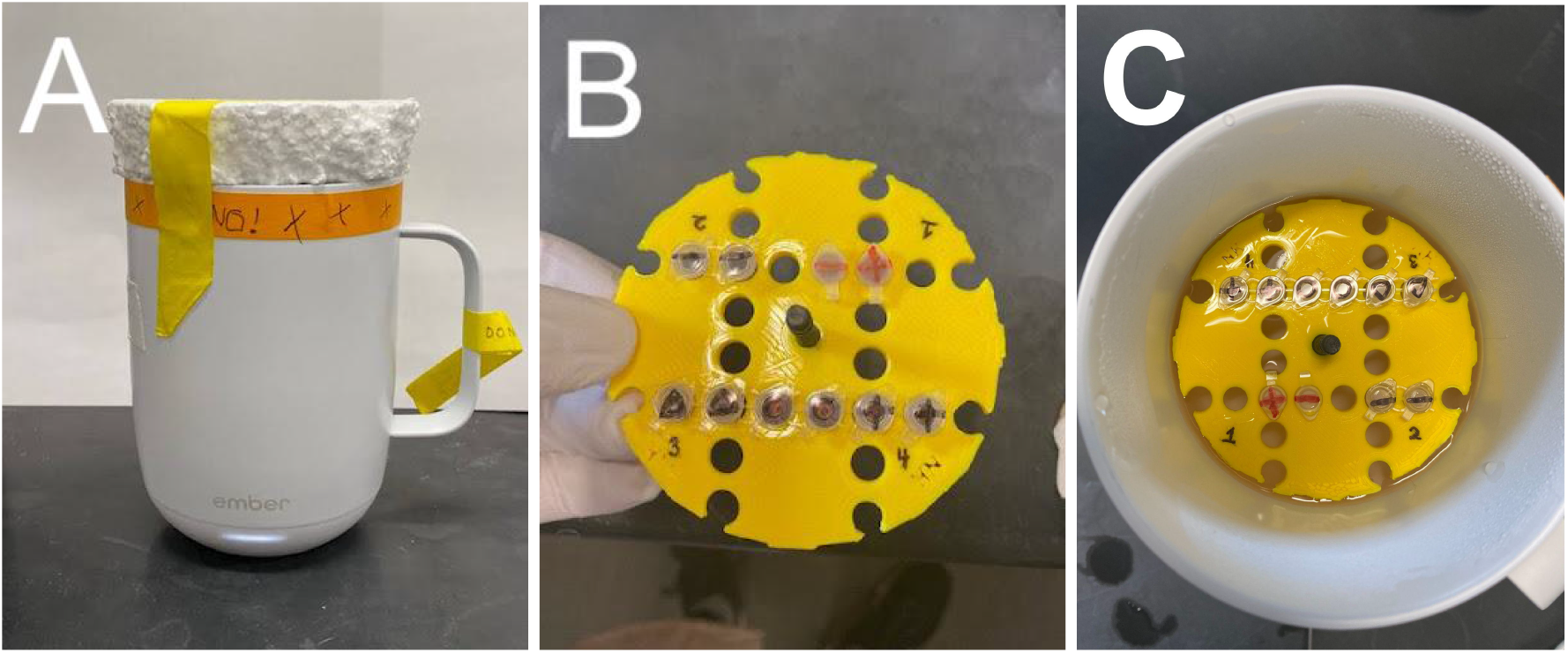
Using the EM2 battery-powered coffee mug heater to incubate the LAMP reactions. The assembled EM2 LAMP diagnostic for HPV-16 using a styrofoam cap to prevent heat loss (A). The reactions were incubated using a 3D-printed float (B) as shown (C).

Using oral rinse as the sample matrix, PTCs were inoculated with 10^4^, 10^3^, or 10^2^ copies of HPV16 E7/E1 DNA to determine the limit of detection (LOD) of the system (Table 6, Figure 4).

**Table 6:** Testing oral rinse as the sample matrix while incubated in the EM2 mug. PTC oral rinse reactions spiked with 10^4^, 10^3^, or 10^2^ copies of HPV16 E7/E1 DNA and NTC oral rinse reactions with no added DNA were scored (P represents no DNA amplification, O represents some DNA amplification, Y represents full DNA amplification, and color combinations represent intermediate levels) every 10 minutes of incubation in an EM2 mug at 65 °C.

|  | 10 min | 20 min | 30 min | 40 min | 50 min | 60 min | 70 min |
| --- | --- | --- | --- | --- | --- | --- | --- |
| PTC $10^4$ | P | O | Y/O | Y | Y | Y | Y |
| PTC $10^4$ | P | O | Y/O | Y | Y | Y | Y |
| PTC $10^3$ | P | O | Y/O | Y | Y | Y | Y |
| PTC $10^3$ | P | O | Y/O | Y | Y | Y | Y |
| PTC $10^2$ | P | P | P | P | P | P | O |
| PTC $10^2$ | P | P | P | P | P | P | O |
| NTC | P | P | P | P | P | P | P/O |
| NTC | P | P | P | P | P | P | P |

**Figure 4:**
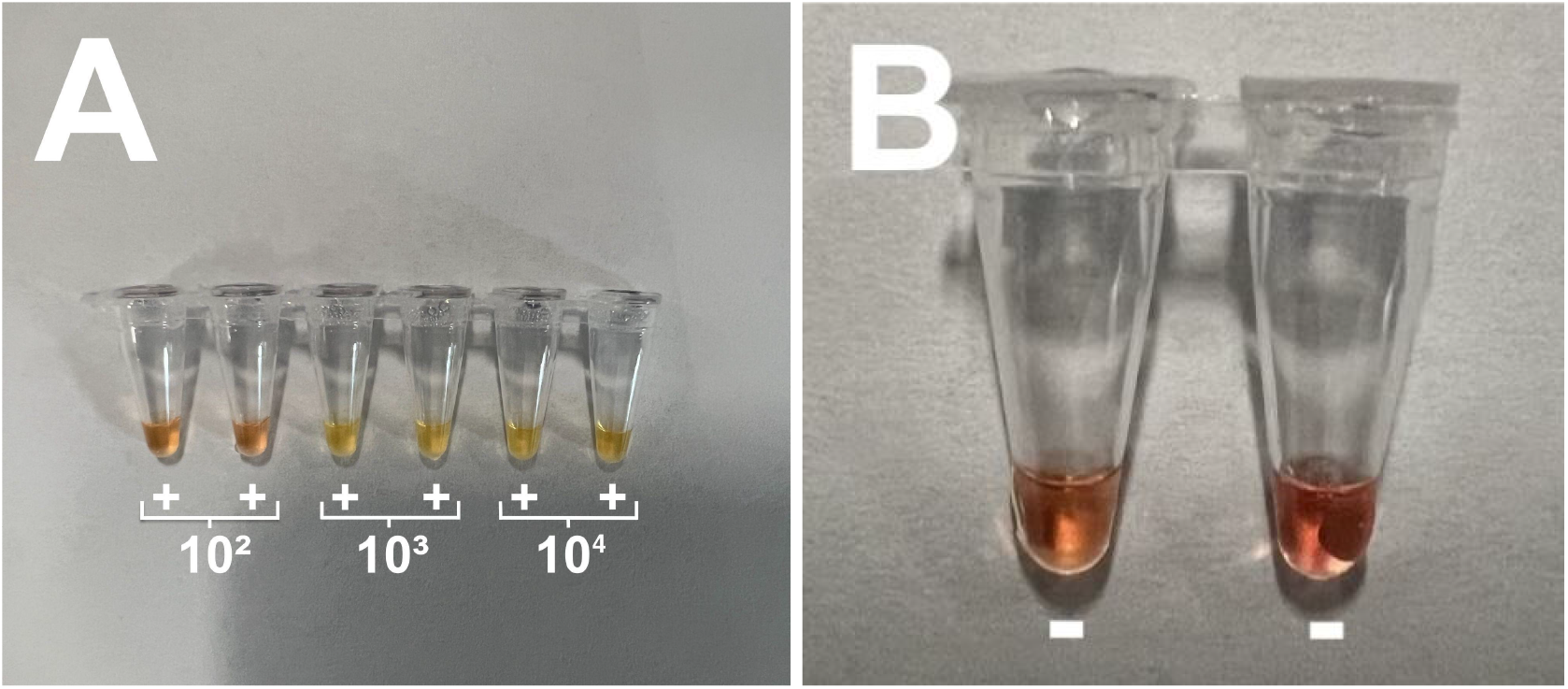
Testing oral rinse as the sample matrix while incubated in the EM2 mug. PTC oral rinse reactions spiked with 10^4^, 10^3^, or 10^2^ copies of HPV16 E7/E1 DNA (A) and NTC oral rinse reactions with no added DNA (B) after 70 minutes of incubation in an EM2 mug at 62.5 °C.

The EM2 mug, while connected to its charger, was able to incubate the LAMP reactions at a stable 62.5 °C for the entire 70-minute duration, resulting in the LAMP assay performing similarly to when it was incubated in the water bath at 65 °C (PTC reactions changed color after 20 minutes and 30 minutes, respectively).

## 4. Discussion

In this study we demonstrated that a user-friendly diagnostic test for oropharyngeal HPV-16 can be assembled with colorimetric LAMP reagents, combined with an oral-rinse protocol, and incubated in a coffee mug. We initially validated the Saetiew et al. 2011 LAMP primers with the NEB colorimetric master mix and found noticeable amplification (signaled by color change) in all positive controls after 30 minutes, while no sign of amplification in negative controls was detected for at least 60 minutes (Section 3.1). Next, we moved on to testing the LAMP system with direct saliva input and an oral rinse protocol. The results indicated that minimally processed oral rinse does not interfere with the colorimetric LAMP read-out or amplification. Unprocessed saliva and saliva diluted as much as 1:3 added directly to the system demonstrated non-specific color changes leading us to determine that it is not compatible with the system. The EM2 device performed reliably, not noticeably deviating from its maximum set point of 62.5 °C while incubating the LAMP reactions which performed in a similar way to water bath incubated reactions.

To the best of our knowledge, this is the first study to demonstrate a successful HPV-16 LAMP reaction carried out in a commercially available battery-powered mug. Additionally, these LAMP reactions received their positive signal through an oral rinse sample, demonstrating an ease-of-use design from start to finish (sample input to interpretation). Limitations of this study include a lack of statistical power in the results due to minimal replicates. Additionally, the LAMP system was not characterized for specificity in this study. The Saetiew et al. (2011) HPV-16 primer set was tested in the original paper for cross-reactivity with HPV-18, 39, 45, 51, 52, 56, 58, and 59 DNA extracted from cervical samples by visually identifying turbidity of the LAMP reaction and performing gel electrophoresis to examine the size of the product. We assume that the specificity demonstrated in that study will continue to apply to the system proposed here.

The system demonstrated a high level of sensitivity in this study using synthetic HPV-16 DNA but future work should determine the sensitivity with clinical samples. Finally, using the oral rinse protocol to sample patients will obviously dilute the viral signal. Research by Steinau et al. (2011) determined that an oral rinse protocol outperformed mucosal and tonsil brushings for detecting HPV in their patients. Future work with this system should pay special attention to keeping the total volume of fluid used to prepare the oral rinse to a minimum, therefore keeping the dilution effect to a minimum. Finally, the EM2 incubation system was tested with the portable mug sitting on the energized charging station saucer throughout the experiments. When off the charger, but filled with prewarmed water, the EM2 is able to maintain a temperature of 62.5 ± 0.1 °C for over 80 minutes (data not shown), which is longer than any LAMP test reported in this study.

While many components of this system could use additional testing and optimization, we believe this study demonstrates a clear step on the path to creating a simple, cheap, and easy-to-use diagnostic for oral HPV-16 infection. Additionally, this study suggests the potential of using a mug-incubated colorimetric LAMP system for other diseases that could benefit from ease-of-use. In general, these LAMP diagnostics have the potential to be especially impactful in economically disadvantaged regions where the accessibility of healthcare is significantly reduced.

## Data Availability

All data produced in the present work are contained in the manuscript.

## Abbreviations

LAMP: Loop-mediated AMPlification
PTC: Positive Template Control
NTC: Negative Template Control
LOD: Limit of Detection
molH_2_O: molecular biology grade nuclease free water
EM2: Ember Temperature Control Smart Mug 2

## 6. Acknowledgements

This work was made possible thanks to generous donations from Bob and Cathy O’Rear and New England Biolabs. We would like to acknowledge support from the community of students that make up the DIY Diagnostic FRI Research Lab https://diystream.cns.utexas.edu/ and mentoring from Andrew D. Ellington and Sanchita Bhadra.

